# Prior infection with SARS-CoV-2 boosts and broadens Ad26.COV2.S immunogenicity in a variant dependent manner

**DOI:** 10.1101/2021.07.24.21261037

**Authors:** Roanne Keeton, Simone I. Richardson, Thandeka Moyo-Gwete, Tandile Hermanus, Marius B. Tincho, Ntombi Benede, Nelia P. Manamela, Richard Baguma, Zanele Makhado, Amkele Ngomti, Thopisang Motlou, Mathilda Mennen, Lionel Chinhoyi, Sango Skelem, Hazel Maboreke, Deelan Doolabh, Arash Iranzadeh, Ashley D. Otter, Tim Brooks, Mahdad Noursadeghi, James Moon, Alba Grifoni, Daniela Weiskopf, Alessandro Sette, Jonathan Blackburn, Nei-Yuan Hsiao, Carolyn Williamson, Catherine Riou, Ameena Goga, Nigel Garrett, Linda-Gail Bekker, Glenda Gray, Ntobeko A. B. Ntusi, Penny L. Moore, Wendy A. Burgers

## Abstract

The Johnson and Johnson Ad26.COV2.S single dose vaccine represents an attractive option for COVID-19 vaccination in resource limited countries. We examined the effect of prior infection with different SARS-CoV-2 variants on Ad26.COV2.S immunogenicity. We compared participants who were SARS-CoV-2 naïve with those either infected with the ancestral D614G virus, or infected in the second wave when Beta predominated. Prior infection significantly boosted spike binding antibodies, antibody-dependent cellular cytotoxicity and neutralizing antibodies against D614G, Beta and Delta, however neutralization cross-reactivity varied by wave. Robust CD4 and CD8 T cell responses were induced after vaccination, regardless of prior infection. T cell recognition of variants was largely preserved, apart from some reduction in CD8 recognition of Delta. Thus, Ad26.COV2.S vaccination following infection may result in enhanced protection against COVID-19. The impact of the infecting variant on neutralization breadth after vaccination has implications for the design of second-generation vaccines based on variants of concern.

## Introduction

The Johnson and Johnson Ad26.COV2.S vaccine is a single dose adenovirus 26 vectored vaccine expressing the SARS-CoV-2 Wuhan-1 stabilized spike. A phase 3 clinical trial of Ad26.COV2.S on three continents demonstrated 66% efficacy against moderate disease and 85% protection against severe disease 28 days after vaccination (Sadoff et al., 2021). Moreover, the South African arm of the trial showed similar levels of efficacy despite the emergence of the neutralization resistant SARS-CoV-2 Beta variant. Vaccination with Ad26.COV2.S triggers neutralizing responses that gradually increase in magnitude and breadth, as well as potent Fc effector functions and T cell activity, both of which retain activity against variants of concern (VOC) (Moore et al., 2021; Barouch et al., 2021; Stephenson et al., 2021; Alter et al., 2021).

Prior infection boosts titers of binding and neutralizing antibodies elicited by mRNA vaccines (Manisty et al., 2021; Saadat et al., 2021; Stamatatos et al., 2021; Vanshylla et al., 2021; Wang et al., 2021b). These increased titers conferred the ability to neutralize SARS-CoV-2 VOCs, illustrating that only one dose of these vaccines may be sufficient to protect previously infected individuals. Similarly, a single dose of the BNT162b2 vaccine boosted antibody-dependent cellular cytotoxicity (ADCC) in previously infected individuals, and T cell cross-reactivity was largely retained (Geers et al., 2021; Reynolds et al., 2021; Tauzin et al., 2021). The impact of prior infection on immune responses elicited by vectored vaccines is less well defined (Havervall et al., 2021), as is the impact of the duration between infection and vaccination, or the genotype of the infecting virus.

## Results

South Africa experienced a first wave of infections in mid-2020, dominated by the ancestral SARS-CoV-2 D614G variant. From November 2020 to February 2021, a second wave of infections was dominated by the Beta variant (Tegally et al., 2021; Wibmer et al., 2021; Cele et al., 2021; Wang et al., 2021a). We established an observational study of 400 healthcare workers (HCWs) with serial sampling since the first wave. We studied 60 HCWs who were vaccinated in a Phase 3b implementation trial of single dose Ad26.COV2.S vaccine (Takuva et al., 2021). HCWs were recruited into three groups, namely those never infected with SARS-CoV-2 (n=20), and those with PCR-confirmed infection during the first wave (n=20) or second wave (n=20) (**Figure 1A** and **Supplemental Table S1**). The Beta variant accounted for >90% of infections in the Western Cape in the second wave (**Figure 1A**), making it likely that this variant was responsible for infections in the latter group. Indeed, whole genome sequencing of 8/20 second wave participants confirmed infection with Beta. Serological profiles were generated for each participant by measuring nucleocapsid and spike antibodies since July 2020 (3-8 monthly visits) (**Supplemental Figure S1A-C**). These data confirmed the absence of infection (or re-infection), and the timing of first or second wave infection. We identified one potential vaccine breakthrough infection and one suspected re-infection, both excluded from subsequent analyses.

**Figure 1.**
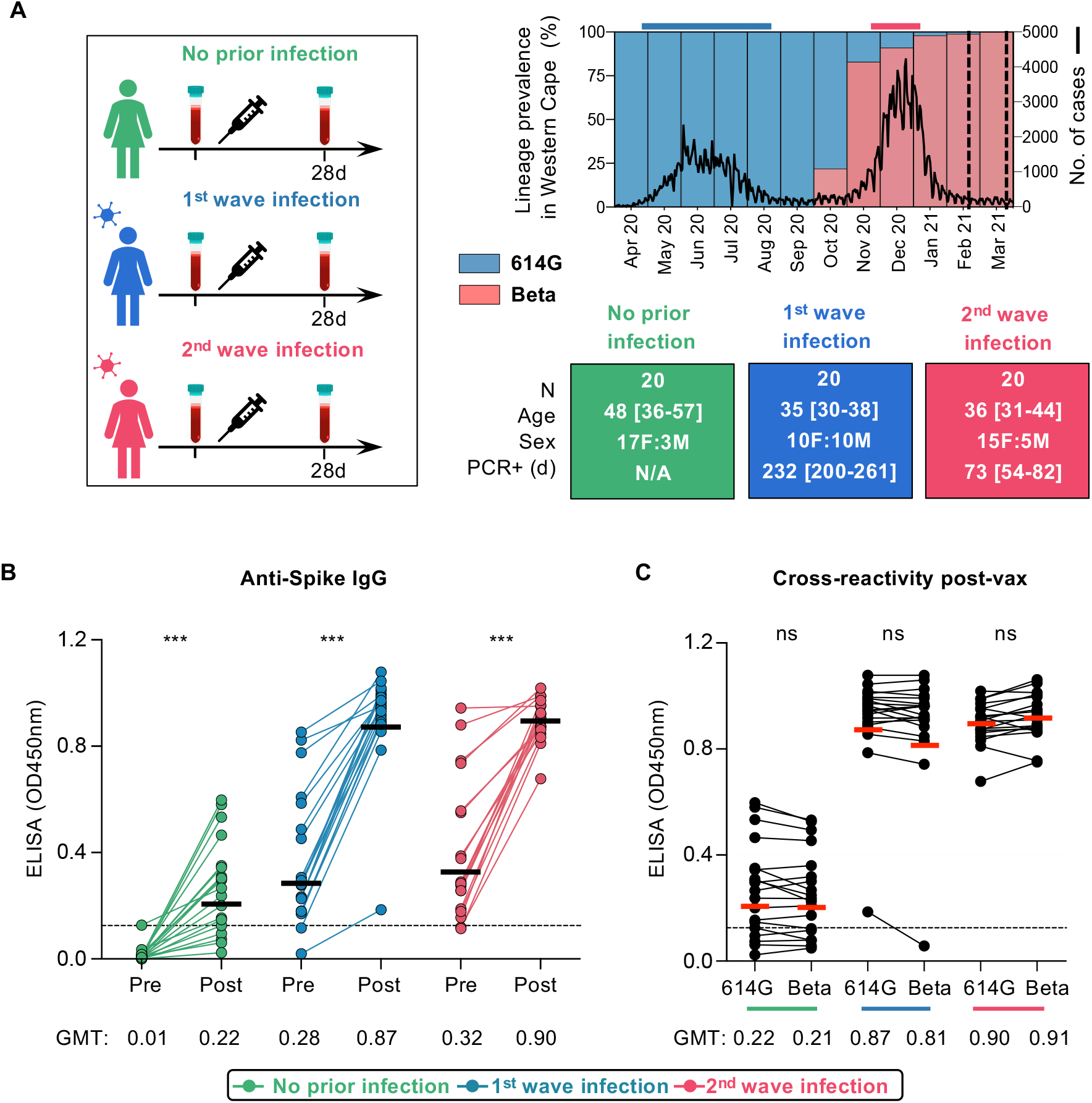
Spike-specific antibody responses in Ad26.COV2.S-vaccinated healthcare workers. **A**. Study design showing three groups (left panel), either with no prior infection, infection in the first wave (May-August 2020) and infection in the second wave (November 2020-January 2021). Samples were taken pre-vaccination and 1 month after vaccination. SARS-CoV-2 epidemiological dynamics in the Western Cape (South Africa) are shown (top right panel). Prevalence of SARS-CoV-2 lineages on the left y-axis. The ancestral strain (D614G) is depicted in blue and Beta in red. The number of COVID-19 cases is represented on the right y-axis. The bars on top of the graph indicate the periods when participants were infected in the first and second waves. Vertical dotted lines indicate when vaccination occurred. Characteristics of participants in the three groups (bottom right panel). Sex, age (median and IQR) and days since PCR-confirmed infection. **B**. Plasma samples from participants with no prior infection (green, n=19), first wave infection (blue, n=20) or second wave infection (red, n=19) were tested for binding to D614G spike protein pre- and post-vaccination (OD_450nm_). **C**. Cross-reactivity of vaccine-induced antibody responses to D614G and Beta spike. The colored lines below the graph correspond with the key. The threshold for positivity is indicated by a dotted line. Horizontal bars indicate GMT, with values shown. Statistical analyses were performed using the Mann-Whitney test between groups, and the Wilcoxon test for pre- and post-vaccine time points or D614G compared to Beta responses. *** denotes p<0.001, ns = non-significant.

Peripheral blood mononuclear cells (PBMC) and plasma were collected prior to vaccination (median 22 days, IQR 14-29) and approximately one month after vaccination (median 29 days, IQR 28-34). We tested pre- and post-vaccination plasma for IgG binding antibodies to the ancestral D614G spike. Binding antibodies elicited by vaccination in the absence of infection (geometric mean titer [GMT]: 0.22) were comparable with those in both infected groups prior to vaccination (GMT: 0.28 and 0.32 for first and second wave, respectively). However, vaccination in HCWs with prior infection in both waves resulted in binding responses being boosted three fold, to a GMT of 0.87 or 0.9 for the first and second waves, respectively (**Figure 1B**). In all HCWs, regardless of prior infection, spike-specific binding antibodies were cross-reactive, with no significant difference in binding between the D614G and Beta spike (**Figure 1C**).

Using a SARS-CoV-2 pseudovirus assay with the D614G spike, we tested neutralizing antibodies elicited by vaccination alone. Consistent with previous studies (Moore et al., 2021), we saw low titers post-vaccination in the infection naïve group (GMT: 74). In both groups with prior infection, we observed a significant boost in neutralization after vaccination against D614G and Beta (**Figure 2A, Supplemental Figure S2A**). For first wave HCWs, titers were boosted 13-fold from a GMT of 210 to 2,798 (**Figure 2A**). Second wave HCWs were boosted 12-fold from a GMT of 99 to 1,157. To determine cross-reactivity of neutralizing antibodies, we compared neutralization of D614G with Beta and Delta. For antibodies induced by vaccination alone, all participants showed significantly lower titers against Beta (85% showing no neutralization, GMT: 28) and Delta (78% showing no neutralization, GMT: 29). In both groups of previously infected HCWs we saw cross-neutralization of Beta and Delta, but the degree of cross-reactivity varied by wave of infection (**Figure 2B and C**). For HCWs infected in the first wave, while neutralization of Beta and Delta was maintained, titers were significantly lower for both VOCs (a reduction in GMT from 2,798 to 606 and 443, respectively, compared to D614G). In contrast, plasma from those infected in the second wave with Beta showed no significant difference in neutralization of D614G (GMT: 1157) but 6-fold lower neutralization of Delta (GMT: 200, p<0.001) (**Figure 2B and C**). Overall, prior infection followed by vaccination triggered high titer neutralizing antibodies able to neutralize VOCs. However, the pattern of neutralization varied by wave, suggesting that the neutralizing antibody repertoire was shaped by the genotype of the infecting variant.

**Figure 2.**
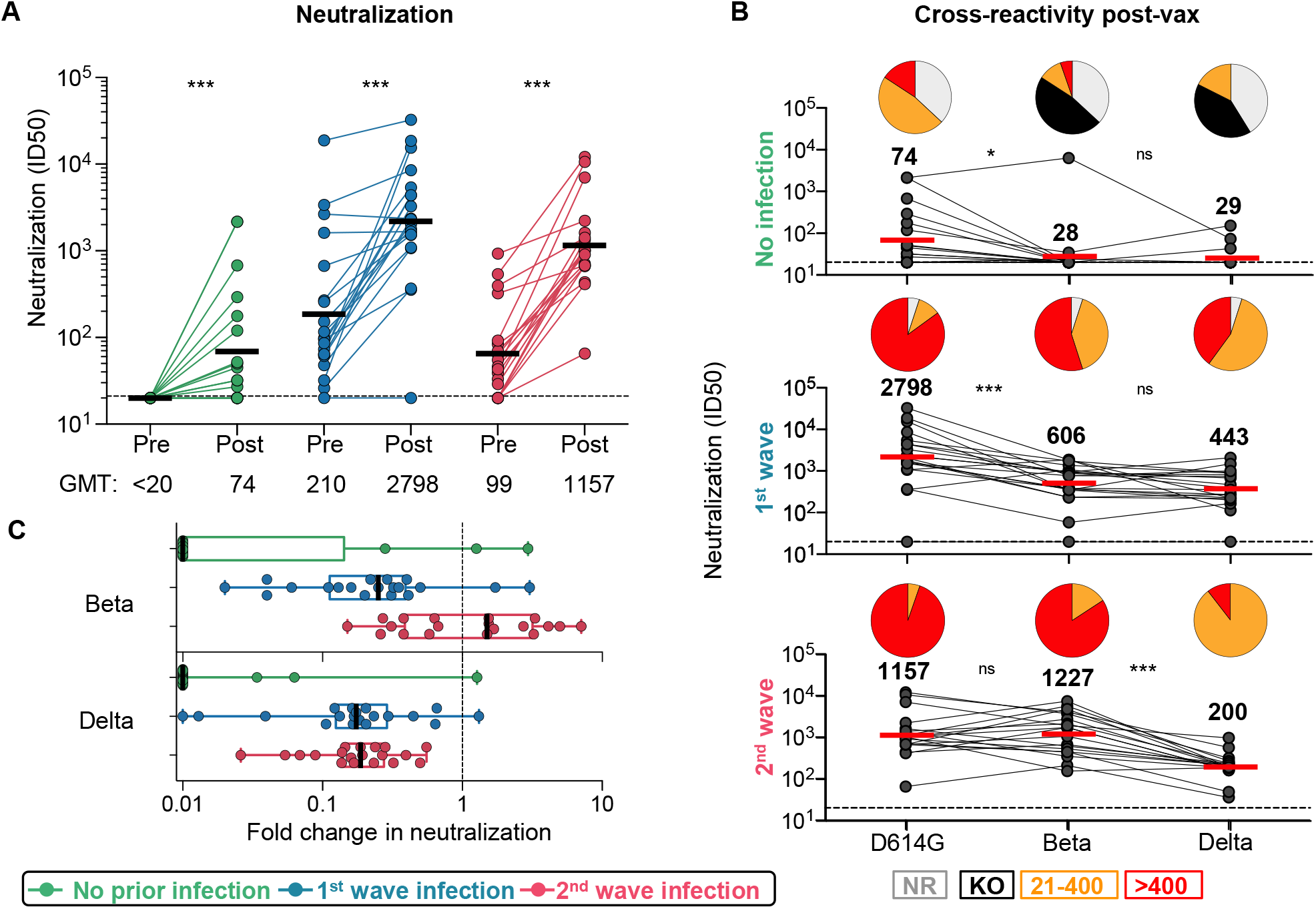
Neutralizing antibody responses to Ad26.COV2.S vaccination. **A**. Neutralization of the SARS-CoV-2 D614G pseudovirus by plasma pre- and post-vaccination from participants with no prior infection (green, n=19) and those infected in the first (blue, n=20) and second waves (red, n=19). Neutralization is reflected as an ID_50_ titer. The threshold for positivity is indicated by a dotted line **B**. Cross-reactive neutralization post-vaccination against D614G, Beta and Delta. Pie charts show the proportion of vaccine non-responders (NR; grey), knock-out of neutralization of Beta or Delta (KO; black), titer of 20-400 (orange), or >400 (red). **C**. Fold change of post-vaccination D614G neutralization titers relative to Beta or Delta. The horizontal bars indicate GMT, with values indicated. Statistical analyses were performed using the Friedman test between groups, and the Wilcoxon test for paired analyses. * denotes p<0.05, *** p<0.001, ns, non-significant.

To assess the impact of prior infection on Fc effector responses to vectored vaccines is unknown. We measured the ability of plasma antibodies to crosslink FcγRIIIa (CD16) expressing cells and cell surface D614G, Beta or Delta spikes on target cells, as a surrogate for ADCC. In previously infected individuals, post-vaccination responses following both waves were significantly higher against D614G, Beta and Delta (**Figure 3A, Supplemental Figure S2B)**, closely mirroring the fold increases of spike binding titers **(Figure 1B)**. However, responses to D614G elicited by vaccination alone (GMT: 39) were similar to those elicited by infection (GMT: 86 for wave 1 and 54 for wave 2) (**Figure 3A, Supplemental Figure S2B)**. ADCC assays performed using the Beta and Delta variants showed no significant loss in activity compared to D614G in the vaccine-only group (**Figure 3B**) or in individuals with prior infection (**Figure 3B and C)**, demonstrating cross-reactive ADCC responses to VOCs.

**Figure 3.**
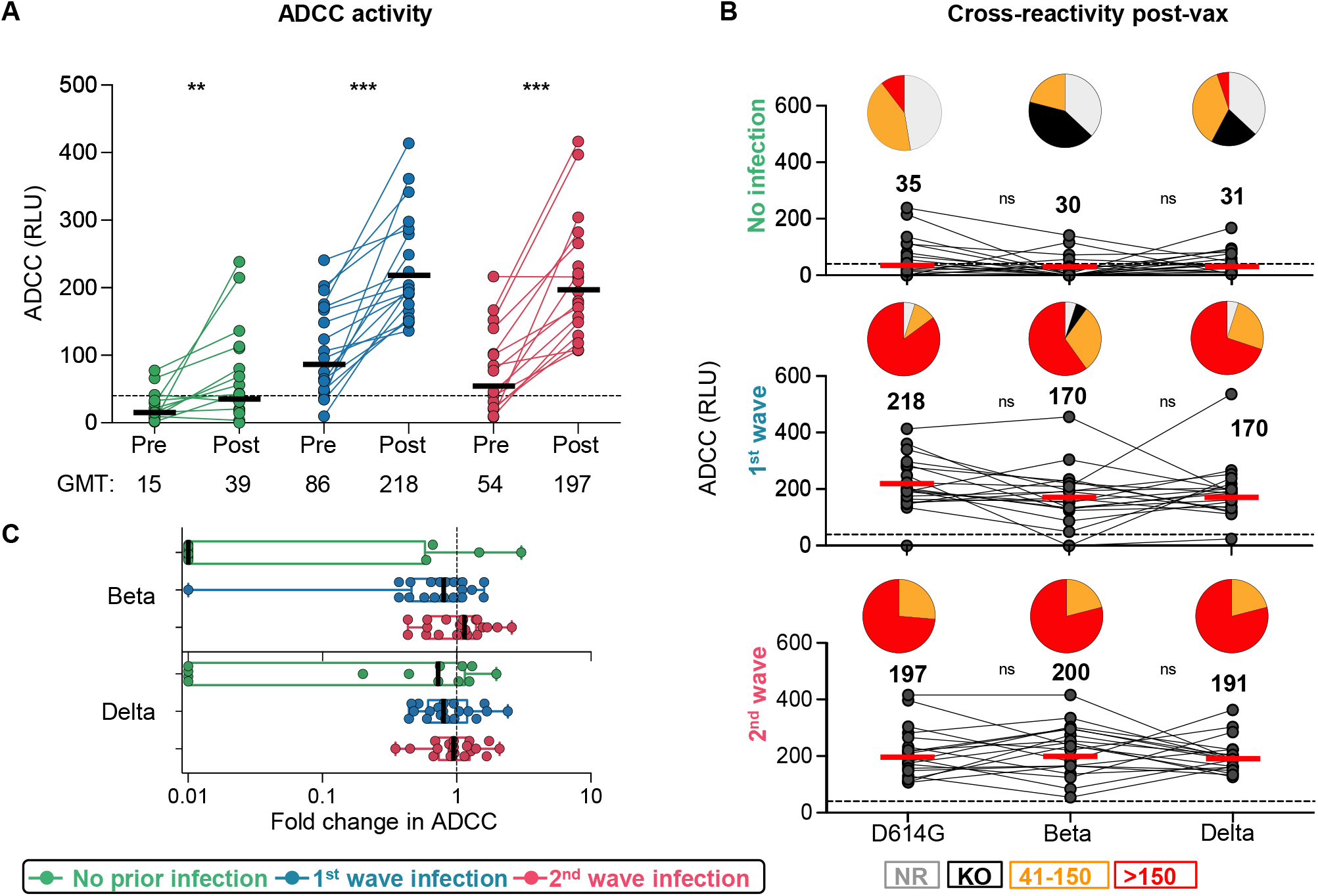
Antibody-dependent cellular cytotoxicity (ADCC) responses to Ad26.COV2.S vaccination. **A**. ADCC activity represented as relative light units (RLU). **B**. Cross-reactive ADCC activity 28 days post-vaccination against D614G, Beta and Delta. Pie charts show the proportion of vaccine non-responders (NR; grey), knock-out of Beta/Delta neutralization (KO; black), or detectable ADCC activity (41-150, orange and >150, red). **C**. Fold-change of post-vaccination D614G ADCC levels relative to the Beta/Delta variants. The horizontal bars indicate GMT, with values indicated. Statistical analyses were performed using the Friedman test between groups, and the Wilcoxon test for pre- and post-vaccine time points or D614G compared to Beta/Delta responses. * denotes p<0.05, ** p<0.01, *** p<0.001, ns, non-significant.

We examined the effect of prior SARS-CoV-2 infection on vaccine T cell responses. We measured intracellular cytokine production (IFN-*γ*, TNF-*α* and IL-2) in response to peptides covering the Wuhan-1 spike (**Supplemental Figure S3A**). Vaccination induced spike-specific CD4 and CD8 T cell responses in all groups (**Figure 4A and B**). Infection-naive participants or those infected in wave 1 had significantly higher CD4 T cell responses after vaccination (median: 0.051 and 0.064, respectively). The wave 2 group had pre-existing responses that were significantly higher than the first wave baseline infection responses, and mounted a more modest response to vaccination, with similar medians (0.132% and 0.147%, p=ns). CD8 responses were present in fewer individuals prior to vaccination (15 and 32%, vs. 85 and 100% for CD4 responses in first and second wave groups, respectively), and did not differ significantly between the groups (**Figure 4B, Supplemental Figure S3B**). Median CD4 T cell frequencies from pre-to post-vaccination decreased with higher magnitude of pre-existing responses, with a 5.7-fold change in the infection-naïve group, 1.5-fold in the first wave group and 1.1-fold in the second wave group. For CD8 responses, the fold-increase was similar for the three groups (**Figure 4C**). A greater proportion of individuals mounted a CD4 response compared to a CD8 response (**Figure 4C**). Polyfunctional profiles of vaccine responses demonstrated that CD4 T cells had the capacity to produce multiple cytokines simultaneously, while CD8 T cells produced predominantly IFN-*γ* alone (**Supplemental Figure S3C**), with no significant difference in the profiles after vaccination for those who were infection naïve compared to those with prior infection.

**Figure 4.**
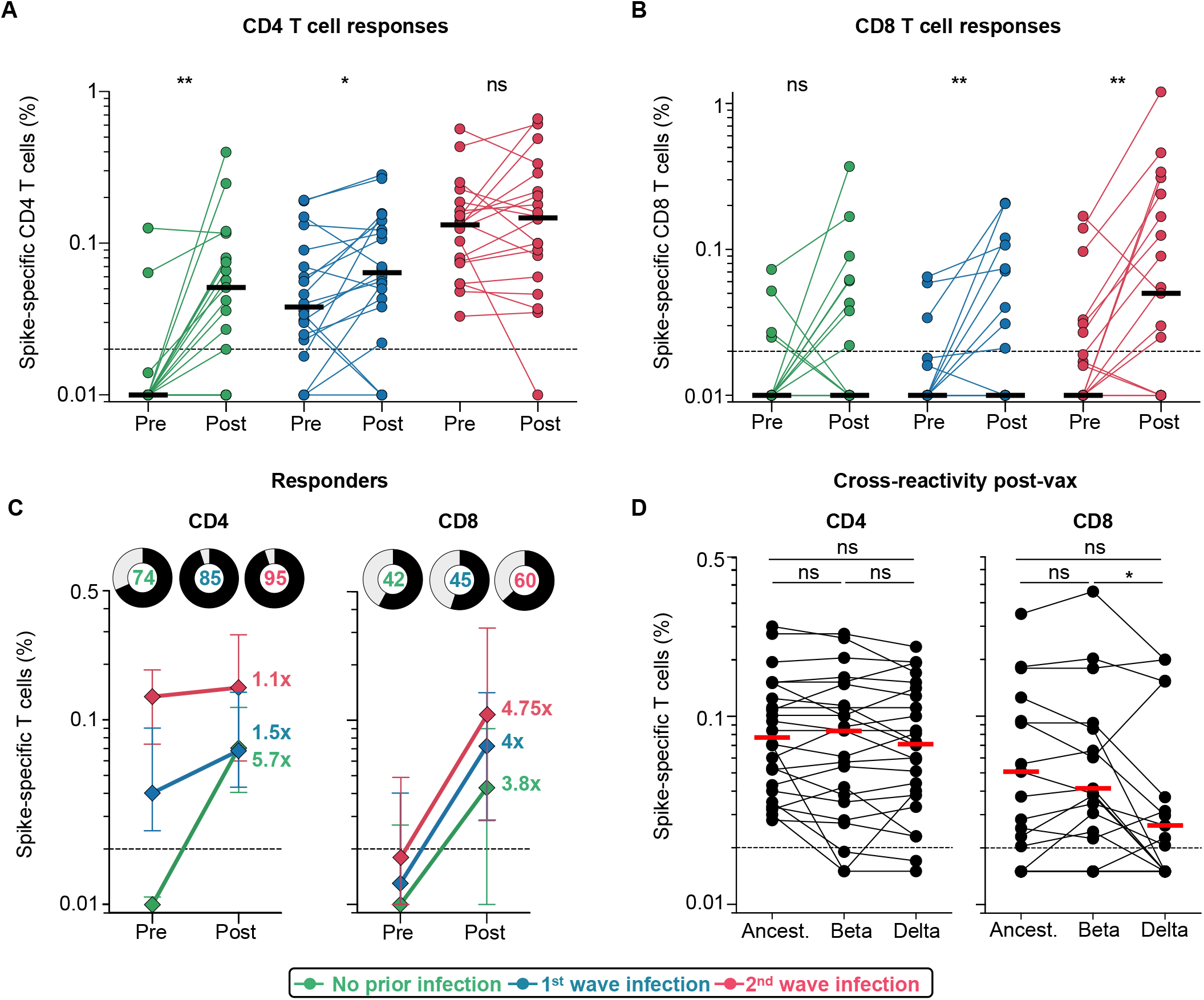
T cell responses to Ad26.COV2.S vaccination. **A**. Frequency of total cytokine-producing spike-specific CD4 T cells, and **B**. CD8 T cells, in those with no prior infection (green, n=19), infection in the first wave (blue, n=20), and infection in the second wave (red, n=19), in PBMC stimulated with peptides based on Wuhan spike. **C**. Median fold-change of CD4 and CD8 T cell frequencies after vaccination in responders. Error bars indicate IQR. Pie charts show responders (black) and non-responders (grey), with the percentage of responders indicated. **D**. Cross-reactivity of T cell responses post-vaccination (n=24) after peptide stimulation with spike from the ancestral strain, Beta or Delta is shown. Horizontal bars indicate medians. The dotted line indicates the threshold for positivity and values are background-subtracted. Statistical analyses were performed using the Wilcoxon test. * denotes p<0.05, ** p<0.01, ns = non-significant.

Finally, we assessed whether T cells induced by vaccination recognized Beta and Delta. We tested spike peptides corresponding to the viral sequences of the ancestral strain, Beta or Delta in 24 vaccinees. CD4 T cell recognition of Beta or Delta was fully preserved compared to the ancestral strain (**Figure 4D**). Spike-specific CD8 T cells (in 15/24 participants) cross-recognised Beta spike in 14/15 responders. In contrast, the median magnitude of the cytokine response was significantly lower against Delta compared to Beta (p=0.041), and 8/15 (53%) of CD8 responders had a two-fold or greater reduction in the response to Delta, including five with complete loss of recognition (**Figure 4D**). Overall, these results demonstrate that robust CD4 and CD8 T cell responses are generated after vaccination, regardless of prior infection. Vaccine T cell cross-recognition of variants is largely preserved, with the exception of a reduced ability for CD8 recognition of Delta in some vaccinees.

## Discussion

Several mRNA vaccines have demonstrated a boosting effect of prior infection (Manisty et al., 2021; Reynolds et al., 2021; Saadat et al., 2021; Stamatatos et al., 2021; Wang et al., 2021b). However, whether this is true of viral vectors, including the single dose Johnson and Johnson vaccine is unclear. We show that infection prior to vaccination with Ad26.COV2.S significantly boosts the magnitude and cross-reactivity of binding antibodies, neutralizing antibodies and Fc effector function. T cell responses were robustly generated even in the absence of prior infection and were preserved against Beta. These data have particular significance in countries like South Africa, where SARS-CoV-2 seropositivity is 20-40% (Mutevedzi et al., 2021; Hsiao et al., 2020; Sykes et al., 2021). Thus, prior infection may enhance the protective efficacy of this vaccine, which is frequently used in resource-limited settings.

Neutralization breadth was shaped by the variant responsible for infection. Prior exposure to D614G resulted in reduced titers against both Beta and Delta, consistent with previous studies (Liu et al., 2021). However, while Beta infection resulted in the preserved neutralization of D614G, we, like others (Liu et al., 2021), noted significant loss of activity against Delta. Therefore, while all participants were exposed to the same vaccine, the genotype of the infecting virus determined the specificity of the responses, prior to vaccine boosting. These findings have important implications for vaccine design as the sequence of VOC spikes in second generation vaccines may impact the repertoire of vaccine-induced antibodies.

Fc effector functions are important in vaccine-elicited protection against many viruses (Richardson and Moore, 2021). Reduced SARS-CoV-2 severity/mortality correlates with Fc effector activity (Zohar et al., 2020) and monoclonal antibodies may require Fc function for optimal protection (Winkler et al., 2021; Schafer et al., 2020). We show that Ad26.COV.2 vaccination in SARS-CoV-2 naïve HCWs elicits significant ADCC responses, consistent with previous data (Stephenson et al., 2021). In addition, prior infection significantly enhanced vaccine-elicited ADCC responses, independent of time post-infection, as for the BNT162b2 vaccine (Geers et al., 2021; Tauzin et al., 2021). Finally, unlike neutralizing antibodies, ADCC activity through vaccination alone, or boosted by prior infection, was cross-reactive for Beta and Delta. This is consistent with previous findings, and suggests that ADCC-mediating antibodies target regions of the spike beyond the major neutralization epitopes (Alter et al., 2021).

SARS-CoV-2-specific T cells play a key role in modulating COVID-19 disease severity (Rydyznski Moderbacher et al., 2020) and provide protective immunity in the context of low antibody titers (McMahan et al., 2020). We show that robust spike-specific CD4 and CD8 memory T cell responses were induced by AD26.COV2.S vaccination, consistent with Alter et al (2021). The magnitude of vaccine-induced T cell responses was similar to convalescent responses. The effect of prior infection was distinct from the antibody response, with CD4 responses in the infection naïve group induced to a similar magnitude as the first wave group, with existing CD4 T cells only moderately boosted, if at all in the second wave group. There was an increase in the magnitude and proportion of CD8 responses to spike induced *de novo* after vaccination, compared to infection, similar for the three study groups. Four participants in the infection naïve group displayed spike-specific CD8 T cells prior to vaccination, as described elsewhere, likely through exposure to endemic human coronaviruses (Braun et al., 2020; Grifoni et al, 2020; Mateus et al., 2020). Of note, most CD8 responses declined after vaccination, suggesting a lack of cognate cross-reactivity.

Vaccination induced T cells that largely cross-recognized peptides based on Beta and Delta spike, suggesting that most vaccinees target conserved epitopes in spike, as previously described (Reynolds et al., 2021; Riou et al., 2021b; Gallagher et al., 2021; Geers et al, 2021; Tarke et al., 2021). However, a third of vaccinees showed reduced CD8 recognition of Delta, which harbours the L452R mutation that confers resistance to HLA-A*24:02 recognition (Motozono et al., 2021). Thus, reduced CD8 recognition of Delta may be attributed to the HLA repertoire of individuals.

Overall, we show a dramatic effect of recent or distant infection on the magnitude and breadth of neutralizing responses and ADCC. Ad26.COV2.S vaccination alone drives continued maturation of B cell responses, conferring enhanced neutralization of variants, and durability (Barouch et al., 2021). Whether prior infection will enhance maturation of neutralizing antibodies and extend durability still further is unknown. T cell responses, though more modestly impacted by prior infection, were robust and largely cross-reactive. This suggests that an infection ‘prime’ boosts Ad26.COV2.S immunogenicity, and in areas of high seroprevalence, may positively impact the effectiveness of this single dose vaccine. Most significantly, we show for the first time that breadth of neutralization after vaccination is dictated by the infecting variant, with important implications for adapted vaccines based on VOCs.

### Limitations of the Study

This study focused on variant-specific responses post-vaccination, and we did not fully assess neutralizing breadth prior to vaccination. However, several studies have now characterised the neutralizing activity against VOCs, including Delta which now dominates globally, and show approximately four-to six-fold reduced sensitivity to convalescent plasma, compared to D614G (Edara et al., 2021; Liu et al, 2021; Planas et al., 2021). Furthermore, whether vectored vaccines predominantly elicit or boost pre-existing RBD responses, like mRNA vaccines (Stamatatos et al., 2021), remains to be defined. Finally, further analyses to identify the HLA alleles associated with reduced CD8 T cell cross-reactivity to Delta, and confirmation of the mutation(s) that are responsible for epitope loss, would shed light on the significance of cellular immune evasion by Delta.

## Data Availability

All data referred to in the manuscript is available in the manuscript or Supplementary files.

## Acknowledgements

We thank the study participants, and the clinical staff and personnel at Groote Schuur Hospital, Cape Town for their dedication. We thank F. Ayres, D. Mhlanga, B. Oosthuysen and B. Lambson for production of protein and pseudoviruses. The parental soluble spike was provided by J. McLellan. The parental pseudovirus plasmids were kindly provided by Drs E. Landais and D. Sok. We thank the Variant consortium of South African scientists. The graphical abstract was created with BioRender.com. This research was supported by the South African Medical Research Council with funds received from the South African Department of Science and Innovation (DSI), including grants 96825, SHIPNCD 76756 and DST/CON 0250/2012. This work was supported by the Poliomyelitis Research Foundation (21/65) and the Wellcome Centre for Infectious Diseases Research in Africa (CIDRI-Africa), which is supported by core funding from the Wellcome Trust (203135/Z/16/Z and 222754). Funding support was received from NIH NIAID with the SARS-CoV-2 Assessment of Viral Evolution program and contract no. 75N9301900065 to A.S. and D.W. P.L.M. and S.I.R. are supported by the South African Research Chairs Initiative of DSI and the National Research Foundation (NRF; No 98341). S.I.R. is a L’Oreal/UNESCO Women in Science South Africa Young Talents awardee. W.A.B. and C.R. are supported by the EDCTP2 programme of the European Union’s Horizon 2020 programme (TMA2017SF-1951-TB-SPEC and TMA2016SF-1535-CaTCH-22). N.A.B.N acknowledges funding from the SA-MRC, MRC UK, NRF and the Lily and Ernst Hausmann Trust. M.N. is supported by the Wellcome Trust (207511/Z/17/Z) and by NIHR Biomedical Research Funding to University College London Hospitals. For the purposes of open access, the authors have applied a CC BY public copyright license to any author-accepted version arising from this submission.

## Author contributions

W.A.B, P.L.M. and N.A.B.N. designed the study. W.A.B and P.L.M. analyzed the data and wrote the manuscript. R.K., S.I.R. and T.M.G. generated and analyzed the data and wrote the manuscript. S.I.R., T.M.G., T.H., N.P.M., Z.M. and T.M. performed antibody assays. R.K., M.B.T., N.B., R.B. and A.N. performed T cell assays. M.M., S.S. and L.R.C. managed the HCW cohort and contributed clinical samples. A.O. and T.B. characterized the serological profiles. N.Y.H. contributed samples and A.G., D.W. and A.S. provided variant peptide pools. D.D., A.I. and C.W. performed viral sequencing. H.M. and J.B. contributed to cohort characterization. C.R. contributed to data analysis. A.G., N.G., L.G.B. and G.G. established and led the Sisonke vaccine study. N.A.B.N., J.M. and M.N. established and led the HCW cohort. All authors reviewed and edited the manuscript.

## STAR METHODS

### RESOURCE AVAILABILITY

#### >Lead contact

Further information and requests for resources and reagents should be directed to and will be fulfilled by the lead contact, Wendy Burgers.

#### Materials availability

Materials will be made available by request to Wendy Burgers.

#### Data and code availability

The published article includes all data generated or analyzed during this study, and summarized in the accompanying tables, figures and supplemental materials.

## EXPERIMENTAL MODEL AND SUBJECT DETAILS

### Human Subjects

Participants were recruited from a longitudinal study of healthcare workers (HCW; n=400) enrolled from Groote Schuur Hospital (Cape Town, Western Cape, South Africa). HCW in this cohort were recruited between July 2020 and January 2021, and vaccination with single dose Johnson and Johnson Ad26.COV2.S in the Sisonke Phase 3b trial took place between 17 February and 26 March 2021. Sixty participants were selected for inclusion in this study, based on the availability of PBMC and plasma prior to vaccination and approximately one month after vaccination, and who fell into one of three groups: (1) No evidence of previous SARS-CoV-2 infection by diagnostic PCR test or serial serology; (2) infection during the ‘first wave’ of the pandemic in South Africa, prior to 1 September 2020, with known date of laboratory (PCR)-confirmed SARS-CoV-2 infection; and (3) infection during the ‘second wave’, with known date of laboratory (PCR)-confirmed SARS-CoV-2 infection between 1 November 2020 and 31 January 2021. Full demographic and clinical characteristics of participants are summarized in **Supplemental Table S1**. The study was approved by the University of Cape Town Human Research Ethics Committee (HREC 190/2020 and 209/2020) and the University of the Witwatersrand Human Research Ethics Committee (Medical) (no M210429). Written informed consent was obtained from all participants.

### Cell Lines

Human embryo kidney HEK293T cells were cultured at 37°C, 5% CO_2_, in DMEM containing 10% heat-inactivated fetal bovine serum (Gibco BRL Life Technologies) and supplemented with 50 μg/ml gentamicin (Sigma). Cells were disrupted at confluence with 0.25% trypsin in 1 mM EDTA (Sigma) every 48–72 hours. HEK293T-ACE2 cells were maintained in the same way as HEK293T cells but were supplemented with 3 μg/ml puromycin for selection of stably transduced cells. Jurkat-Lucia™ NFAT-CD16 cells were maintained in IMDM media with 10% heat-inactivated fetal bovine serum (Gibco, Gaithersburg, MD), 1% Penicillin Streptomycin (Gibco, Gaithersburg, MD) and 10 μg/ml of Blasticidin and 100 μg/ml of Zeocin was added to the growth medium every other passage.

## METHOD DETAILS

### SARS-CoV-2 spike WGS and phylogenetic analysis

Whole genome sequencing (WGS) of SARS-CoV-2 was performed using nasopharyngeal swabs obtained from 19 of the hospitalized patients recruited during the second COVID-19 wave. Sequencing was performed as previously published (Moyo-Gwete et al., 2021). Briefly, cDNA was synthesized from RNA extracted from the nasopharyngeal swabs using the Superscript IV First Strand synthesis system (Life Technologies, Carlsbad, CA) and random hexamer primers. Whole genome amplification was then performed by multiplex PCR using the ARTIC V3 protocol (https://www.protocols.io/view/ncov-2019-sequencing-protocol-v3-locost-bh42j8ye). PCR products were purified with AMPure XP magnetic beads (Beckman Coulter, CA) and quantified using the Qubit dsDNA High Sensitivity assay on the Qubit 3.0 instrument (Life Technologies Carlsbad, CA). The Illumina® DNA Prep kit was used to prepare indexed paired end libraries of genomic DNA. Sequencing libraries were normalized to 4 nM, pooled, and denatured with 0.2 N sodium hydroxide. Libraries were sequenced on the Illumina MiSeq instrument (Illumina, San Diego, CA, USA). The quality control checks on raw sequence data and the genome assembly were performed using Genome Detective 1.132 (https://www.genomedetective.com) and the Coronavirus Typing Tool (Cleemput et al., 2020). The initial assembly obtained from Genome Detective was polished by aligning mapped reads to the references and filtering out low-quality mutations using bcftools 1.7-2 mpileup method. Mutations were confirmed visually with bam files using Geneious software (Biomatters Ltd, New Zealand). Phylogenetic clade classification of the genomes in this study consisted of analyzing them against a global reference dataset using a custom pipeline based on a local version of NextStrain (https://github.com/nextstrain/ncov) (Hadfield et al., 2018).

### Roche serology

Serial serum samples were analysed from longitudinal study visits from enrolment to post-vaccination (3-8 time points per participant) at Public Health England, Porton Down. The Elecsys anti-SARS-CoV-2 Spike and the Elecys anti-SARS-CoV-2 electrochemiluminescent immunoassays were performed (Roche Diagnostics, GmbH), which enable detection of total antibodies against the SARS-CoV-2 spike (S) receptor binding domain (RBD) and nucleocapsid (N) proteins, respectively. Samples were analysed on a Cobas e801 instrument and a result ≥0.8 U/mL was considered positive in the S assay, and ≥1.0 U/mL positive in the N assay, according to the manufacturer’s instructions.

### Isolation of PBMC

Blood was collected in heparin tubes and processed within 3 hours of collection. Peripheral blood mononuclear cells (PBMC) were isolated by density gradient sedimentation using Ficoll-Paque (Amersham Biosciences, Little Chalfont, UK) as per the manufacturer’s instructions and cryopreserved in freezing media consisting of heat-inactivated fetal bovine serum (FBS, Thermofisher Scientific) containing 10% DMSO and stored in liquid nitrogen until use.

### SARS-CoV-2 antigens

For serology assays, SARS-CoV-2 original and beta variant spike proteins were expressed in Human Embryonic Kidney (HEK) 293F suspension cells by transfecting the cells with the spike plasmid. After incubating for six days at 37°C, 70% humidity and 10% CO_2_, proteins were first purified using a nickel resin followed by size-exclusion chromatography. Relevant fractions were collected and frozen at -80 °C until use.

For T cell assays, we used peptides covering the full length SARS-CoV-2 spike protein, by combining two commercially available peptide pools of 15mer sequences with 11 amino acids (aa) overlap (PepTivator®, Miltenyi Biotech, Bergisch Gladbach, Germany). These peptides are based on the Wuhan-1 strain and cover the N-terminal S1 domain of SARS-CoV-2 from aa 1 to 692, as well as the majority of the C-terminal S2 domain. Pools were resuspended in distilled water at a concentration of 50 μg/mL and used at a final concentration of 1 μg/mL. To determine T cell responses to SARS-CoV-2 variants, peptides were synthesized that spanned the entire SARS-CoV-2 spike protein and corresponded to the ancestral Wuhan sequence (GenBank: MN908947) or the Beta (B.1.351; GISAID: EPI_ISL_736932, EPI_ISL_736944, EPI_ISL_736971, EPI_ISL_736966, EPI_ISL_736973, EPI_ISL_825104, EPI_ISL_825120, EPI_ISL_825131), as previously reported (Tarke et al., 2021) or to the Delta SARS-CoV-2 variants (B.1.617.2; GISAID: EPI_ISL_2020950). Peptides were 15-mers overlapping by 10 amino acids and were synthesized as crude material (TC Peptide Lab, San Diego, CA). All peptides were individually resuspended in dimethyl sulfoxide (DMSO) at a concentration of 10–20 mg/mL. Megapools for each antigen were created by pooling aliquots of these individual peptides in the respective SARS-CoV-2 spike sequences, followed by sequential lyophilization steps, and resuspension in DMSO at 1 mg/mL. Pools were used at a final concentration of 1 μg/mL with an equimolar DMSO concentration in the non-stimulated control.

### SARS-CoV-2 Spike ELISA

Two μg/ml of spike protein were used to coat 96-well, high-binding plates and incubated overnight at 4°C. The plates were incubated in a blocking buffer consisting of 5% skimmed milk powder, 0.05% Tween 20, 1x PBS. Plasma samples were diluted to 1:100 starting dilution in a blocking buffer and added to the plates. Secondary antibody was diluted to 1:3000 in blocking buffer and added to the plates followed by TMB substrate (Thermofisher Scientific). Upon stopping the reaction with 1 M H_2_SO_4_, absorbance was measured at a 450nm wavelength. In all instances, mAbs CR3022 and BD23 were used as positive controls and palivizumab was used as a negative control. All values were normalized with the CR3022 mAb.

### Pseudovirus neutralization assay

SARS-CoV-2 pseudotyped lentiviruses were prepared by co-transfecting the HEK 293T cell line with either the SARS-CoV-2 ancestral variant spike (D614G), the Beta spike (L18F, D80A, D215G, K417N, E484K, N501Y, D614G, A701V, 242-244 del) or the Delta spike (T19R, R158G L452R, T478K, D614G, P681R, D950N, 156-157 del) plasmids in conjunction with a firefly luciferase encoding lentivirus backbone plasmid. For the neutralization assay, heat-inactivated plasma samples from vaccine recipients were incubated with the SARS-CoV-2 pseudotyped virus for 1 hour at 37°C, 5% CO_2_. Subsequently, 1×104 HEK 293T cells engineered to over-express ACE-2 were added and incubated at 37°C, 5% CO_2_ for 72 hours upon which the luminescence of the luciferase gene was measured. CB6 was used as a positive control.

### ADCC assay

The ability of plasma antibodies to cross-link FcγRIIIa (CD16) and spike expressing cells was measured as a proxy for antibody-dependent cellular cytotoxicity (ADCC). HEK293T cells were transfected with 5μg of SARS-CoV-2 original variant spike (D614G), Beta or Delta spike plasmids using PEI-MAX 40,000 (Polysciences) and incubated for 2 days at 37°C. Expression of spike was confirmed by binding of CR3022 and P2B-2F6 and their detection by anti-IgG APC staining measured by flow cytometry. Subsequently, 1×10^5^ spike transfected cells per well were incubated with heat inactivated plasma (1:100 final dilution) or control mAbs (final concentration of 100 μg/ml) in RPMI 1640 media supplemented with 10% FBS 1% Pen/Strep (Gibco, Gaithersburg, MD) for 1 hour at 37°C. Jurkat-Lucia™ NFAT-CD16 cells (Invivogen) (2×10^5^ cells/well) were added and incubated for 24 hours at 37°C, 5% CO_2_. Twenty μl of supernatant was then transferred to a white 96-well plate with 50 μl of reconstituted QUANTI-Luc secreted luciferase and read immediately on a Victor 3 luminometer with 1s integration time. Relative light units (RLU) of a no antibody control were subtracted as background. Palivizumab was used as a negative control, while CR3022 was used as a positive control, and P2B-2F6 to differentiate the Beta from the D614G variant. To induce the transgene 1x cell stimulation cocktail (Thermofisher Scientific, Oslo, Norway) and 2 μg/ml ionomycin in R10 was added as a positive control.

### Cell stimulation and flow cytometry staining

Cryopreserved PBMC were thawed, washed and rested in RPMI 1640 containing 10% heat-inactivated FCS for 4 hours prior to stimulation. PBMC were seeded in a 96-well V-bottom plate at ∼2 × 10^6^ PBMC per well and stimulated with SARS-CoV-2 spike peptide pools: full spike pool (Miltenyi), and ancestral and beta mutated S1 and S2 pools (1 μg/mL). All stimulations were performed in the presence of Brefeldin A (10 μg/mL, Sigma-Aldrich, St Louis, MO, USA) and co-stimulatory antibodies against CD28 (clone 28.2) and CD49d (clone L25) (1 μg/mL each; BD Biosciences, San Jose, CA, USA). As a negative control, PBMC were incubated with co-stimulatory antibodies, Brefeldin A and an equimolar amount of DMSO.

After 16 hours of stimulation, cells were washed, stained with LIVE/DEAD™ Fixable VIVID Stain (Invitrogen, Carlsbad, CA, USA) and subsequently surface stained with the following antibodies: CD14 Pac Blue (TuK4, Invitrogen Thermofisher Scientific), CD19 Pac Blue (SJ25-C1, Invitrogen Thermofisher Scientific), CD4 PERCP-Cy5.5 (L200, BD Biosciences, San Jose, CA, USA), CD8 BV510 (RPA-8, Biolegend, San Diego, CA, USA), PD-1 BV711 (EH12.2H7, Biolegend, San Diego, CA, USA), CD27 PE-Cy5 (1A4, Beckman Coulter), CD45RA BV570 (HI100, Biolegend, San Diego, CA, USA). Cells were then fixed and permeabilized using a Cytofix/Cyto perm buffer (BD Biosciences) and stained with CD3 BV650 (OKT3) IFN-g Alexo 700 (B27), TNF BV786 (Mab11) and IL-2 APC (MQ1-17H12) from Biolegend. Finally, cells were washed and fixed in CellFIX (BD Biosciences). Samples were acquired on a BD LSR-II flow cytometer and analyzed using FlowJo (v10, FlowJo LLC, Ashland, OR, USA). A median of 282 848 CD4 events (IQR:216 796 - 355 414) and 153 192 CD8 events (IQR 109 697 - 202 204) were acquired. Cells were gated on singlets, CD14-CD19-, live lymphocytes and memory cells (excluding naive CD27+ CD45RA+ population). Results are expressed as the frequency of CD4+ or CD8+ T cells expressing IFN-g, TNF-a or IL-2. Due to high TNF-a backgrounds, cells producing TNF-a alone were excluded from the analysis. Cytokine responses presented are background subtracted values (from the frequency of cytokine produced in unstimulated cells), and the threshold for a positive cytokine response was defined as >0.02%.

## QUANTIFICATION AND STATISTICAL ANALYSIS

Statistical analyses were performed in Prism (v9; GraphPad Software Inc, San Diego, CA, USA). Non-parametric tests were used for all comparisons. The Mann-Whitney, Friedman and Wilcoxon tests were used for unmatched and paired samples, respectively. All correlations reported are non-parametric Spearman’s correlations. *P* values less than 0.05 were considered statistically significant. Details of analysis performed for each experiment are described in the figure legends.

## References

Alter, G., Yu, J., Liu, J., Chandrashekar, A., Borducchi, E.N., Tostanoski, L.H., McMahan, K., Jacob-Dolan, C., Nkolola, J., Stephenson, K.E., et al. (2021). Immunogenicity of Ad26.COV2.S Against SARS-CoV-2 Variants. Nature 9, 1–9. http://dx.doi.org/10.1038/s41586-021-03681-2.

Anand, S.P., Prevost, J., Nayrac, M., Beaudoin-Bussieres, G., Benlarbi, M., Gasser, R., Brassard, N., Laumaea, A., Gong, S.Y., Bourassa, C., et al. (2021). Longitudinal analysis of humoral immunity against SARS-CoV-2 Spike in convalescent individuals up to 8 months post-symptom onset. Cell Rep Med 2. http://dx.doi.org/10.1016/j.xcrm.2021.100290.

Barouch, D.H., Stephenson, K.E., Sadoff, J., Yu, J., Chang, A., Gebre, M., McMahan, K., Liu, J., Chandrashekar, A., and Patel, S. (2021). Durable Humoral and Cellular Immune Responses 8 Months after Ad26.COV2.S Vaccination. New England Journal of Medicine. http://dx.doi.org/10.1056/NEJMc2108829.

Barrett, J.R., Belij-Rammerstorfer, S., Dold, C., Ewer, K.J., Folegatti, P.M., Gilbride, C., Halkerston, R., Hill, J., Jenkin, D., Stockdale, L., et al. (2021). Phase 1/2 trial of SARS-CoV-2 vaccine ChAdOx1 nCoV-19 with a booster dose induces multifunctional antibody responses. Nat Med 27, 279–288. http://dx.doi.org/10.1038/s41591-020-01179-4.

Braun, J., Loyal, L., Frentsch, M., Wendisch, D., Georg, P., Kurth, F., Hippenstiel, S., Dingeldey, M., Kruse, B., Fauchere, F., et al. (2020). SARS-CoV-2-reactive T cells in healthy donors and patients with COVID-19. Nature 587, 270–274. https://dx.doi.org/10.1038/s41586-020-2598-9.

Brouwer, P.J., Caniels, T.G., van der Straten, K., Snitselaar, J.L., Aldon, Y., Bangaru, S., Torres, J.L., Okba, N.M., Claireaux, M., and Kerster, G. (2020). Potent neutralizing antibodies from COVID-19 patients define multiple targets of vulnerability. Science 369, 643–650. http://dx.doi.org/10.1126/science.abc5902.

Cele, S., Gazy, I., Jackson, L., Hwa, S.H., Tegally, H., Lustig, G., Giandhari, J., Pillay, S., Wilkinson, E., Naidoo, Y., et al. (2021). Escape of SARS-CoV-2 501Y.V2 from neutralization by convalescent plasma. Nature 593, 142–146. http://dx.doi.org/10.1038/s41586-021-03471-w.

Cleemput, S., Dumon, W., Fonseca, V., Abdool Karim, W., Giovanetti, M., Alcantara, L.C., Deforche, K., and De Oliveira, T. (2020). Genome Detective Coronavirus Typing Tool for rapid identification and characterization of novel coronavirus genomes. Bioinformatics 36, 3552–3555. http://dx.doi.org/10.1093/bioinformatics/btaa145.

Edara, V.V., Pinsky, B.A., Suthar, M.S., Lai, L., Davis-Gardner, M.E., Floyd, K., Flowers, M.W., Wrammert, J., Hussaini, L., Ciric, C.R., et al. (2021). Infection and Vaccine-Induced Neutralizing-Antibody Responses to the SARS-CoV-2 B.1.617 Variants. New England Journal of Medicine 12, 664–666. http://dx.doi.org/10.1056/nejmc2107799.

Gallagher, K.M.E., Leick, M.B., Larson, R.C., Berger, T.R., Katsis, K., Yam, J.Y., Brini, G., Grauwet, K., Collection, M.C.-., Processing, T., and Maus, M.V. (2021). SARS -CoV-2 T-cell immunity to variants of concern following vaccination. bioRxiv. http://dx.doi.org/10.1101/2021.05.03.442455.

Geers, D., Shamier, M.C., Bogers, S., den Hartog, G., Gommers, L., Nieuwkoop, N.N., Schmitz, K.S., Rijsbergen, L.C., van Osch, J.A.T., Dijkhuizen, E., et al. (2021). SARS-CoV-2 variants of concern partially escape humoral but not T-cell responses in COVID-19 convalescent donors and vaccinees. Sci Immunol 6. http://dx.doi.org/10.1126/sciimmunol.abj1750.

Grifoni, A., Weiskopf, D., Ramirez, S.I., Mateus, J., Dan, J.M., Moderbacher, C.R., Rawlings, S.A., Sutherland, A., Premkumar, L., Jadi, R.S., et al. (2020). Targets of T Cell Responses to SARS-CoV-2 Coronavirus in Humans with COVID-19 Disease and Unexposed Individuals. Cell 181, 1489-1501.e15. https://dx.doi.org/10.1016/j.cell.2020.05.015

Hadfield, J., Megill, C., Bell, S.M., Huddleston, J., Potter, B., Callender, C., Sagulenko, P., Bedford, T., and Neher, R.A. (2018). Nextstrain: real-time tracking of pathogen evolution. Bioinformatics 34, 4121–4123. http://dx.doi.org/10.1093/bioinformatics/bty407.

Havervall, S., Marking, U., Greilert-Norin, N., Ng, H., Salomonsson, A.-C., Hellström, C., Pin, E., Blom, K., Mangsbo, S., Phillipson, M., et al. (2021). Antibody Responses After a Single Dose of ChAdOx1 nCoV-19 Vaccine in Healthcare Workers Previously Infected with SARS-CoV-2. medRxiv. http://dx.doi.org/10.1101/2021.05.08.21256866.

Hsiao, M., Davies, M., and Kalk, E. (2020). SARS-CoV-2 seroprevalence in the Cape Town Metropolitan sub-districts after the peak of infections. NICD COVID-19 Special Public Health Surveill Bull 18, 1–9.

Klingler, J., Lambert, G.S., Itri, V., Liu, S., Bandres, J.C., Enyindah-Asonye, G., Liu, X., Oguntuyo, K.Y., Amanat, F., and Lee, B. (2021). SARS-CoV-2 mRNA vaccines induce a greater array of spike-specific antibody isotypes with more potent complement binding capacity than natural infection. medRxiv. http://dx.doi.org/10.1101/2021.05.11.21256972.

Lee, W.S., Selva, K.J., Davis, S.K., Wines, B.D., Reynaldi, A., Esterbauer, R., Kelly, H.G., Haycroft, E.R., Tan, H.X., Juno, J.A., et al. (2021). Decay of Fc-dependent antibody functions after mild to moderate COVID-19. Cell Rep Med 2, 100296. http://dx.doi.org/10.1016/j.xcrm.2021.100296.

Liu, C., Ginn, H.M., Dejnirattisai, W., Supasa, P., Wang, B., Tuekprakhon, A., Nutalai, R., Zhou, D., Mentzer, A.J., Zhao, Y., et al. (2021). Reduced neutralization of SARS-CoV-2 B. 1.617 by vaccine and convalescent serum. Cell 184, 1–17. http://dx.doi.org/10.1016/j.cell.2021.06.020.

Manisty, C., Otter, A.D., Treibel, T.A., McKnight, Á., Altmann, D.M., Brooks, T., Noursadeghi, M., Boyton, R.J., Semper, A., and Moon, J.C. (2021). Antibody response to first BNT162b2 dose in previously SARS-CoV-2-infected individuals. The Lancet 397, 1057–1058. http://dx.doi.org/10.1016/S0140-6736(21)00501-8.

Mateus, J., Grifoni, A., Tarke, A., Sidney, J., Ramirez, S.I., Dan, J.M., Burger, Z.C., Rawlings, S.A., Smith, D.M., Phillips, E., et al. (2020). Selective and cross-reactive SARS-CoV-2 T cell epitopes in unexposed humans. Science 370, 89–94. https://dx.doi.org/10.1126/science.abd3871.

McMahan, K., Yu, J., Mercado, N.B., Loos, C., Tostanoski, L.H., Chandrashekar, A., Liu, J., Peter, L., Atyeo, C., Zhu, A., et al. (2021). Correlates of protection against SARS-CoV-2 in rhesus macaques. Nature 590, 630–634. http://dx.doi.org/10.1038/s41586-020-03041-6.

Mercado, N.B., Zahn, R., Wegmann, F., Loos, C., Chandrashekar, A., Yu, J., Liu, J., Peter, L., McMahan, K., and Tostanoski, L.H. (2020). Single-shot Ad26 vaccine protects against SARS-CoV-2 in rhesus macaques. Nature 586, 583–588. http://dx.doi.org/10.1038/s41586-020-2607-z.

Moore, P., Moyo-Gwete, T., Hermanus, T., Kgagudi, P., Ayres, F., Makhado, Z., Sadoff, J., Le Gars, M., van Roey, G., Crowther, C., et al. (2021). Neutralizing antibodies elicited by the Ad26.COV2.S COVID-19 vaccine show reduced activity against 501Y.V2 (B. 1.351), despite protection against severe disease by this variant. bioRxiv. http://dx.doi.org/10.1101/2021.06.09.447722.

Motozono, C., Toyoda, M., Zahradnik, J., Saito, A., Nasser, H., Tan, T.S., Ngare, I., Kimura, I., Uriu, K., Kosugi, Y., et al. (2021). SARS-CoV-2 spike L452R variant evades cellular immunity and increases infectivity. Cell Host & Microbe 29, 1124-1136.e11. https://dx.doi.org/10.1016/j.chom.2021.06.006.

Moyo-Gwete, T., Madzivhandila, M., Makhado, Z., Ayres, F., Mhlanga, D., Oosthuysen, B., Lambson, B.E., Kgagudi, P., Tegally, H., Iranzadeh, A., et al. (2021). Cross-Reactive Neutralizing Antibody Responses Elicited by SARS-CoV-2 501Y.V2 (B.1.351). New England Journal of Medicine. http://dx.doi.org/10.1056/NEJMc2104192.

Mutevedzi, P.C., Kawonga, M., Kwatra, G., Moultrie, A., Baillie, V.L., Mabhena, N., Mathibe, M.N., Rafuma, M.M., Maposa, I., and Abbott, G. (2021). Population Based SARS-CoV-2 Sero-Epidemiological Survey and Estimated Infection Incidence and Fatality Risk in Gauteng Province, South Africa. Available at SSRN: https://ssrn.com/abstract=3855442. http://dx.doi.org/10.2139/ssrn.3855442.

Nielsen, S.C., Yang, F., Jackson, K.J., Hoh, R.A., Röltgen, K., Jean, G.H., Stevens, B.A., Lee, J.-Y., Rustagi, A., and Rogers, A.J. (2020). Human B cell clonal expansion and convergent antibody responses to SARS-CoV-2. Cell Host & Microbe 28, 516-525.e515. http://dx.doi.org/10.1016/j.chom.2020.09.002.

Planas, D., Veyer, D., Baidaliuk, A., Staropoli, I., Guivel-Benhassine, F., Rajah, M.M., Planchais, C., Porrot, F., Robillard, N., Puech, J., et al. (2021). Reduced sensitivity of SARS-CoV-2 variant Delta to antibody neutralization. Nature 596, 276–280. https://dx.doi.org/10.1038/s41586-021-03777-9.

Reynolds, C.J., Pade, C., Gibbons, J.M., Butler, D.K., Otter, A.D., Menacho, K., Fontana, M., Smit, A., Sackville-West, J.E., Cutino-Moguel, T., et al. (2021). Prior SARS-CoV-2 infection rescues B and T cell responses to variants after first vaccine dose. Science 372, 1418–1423. https://dx.doi.org/10.1126/science.abh1282.

Richardson, S.I., and Moore, P.L. (2021). Targeting Fc effector function in vaccine design. Expert Opin Ther Targets, 1–11. http://dx.doi.org/10.1080/14728222.2021.1907343.

Riou, C., du Bruyn, E., Stek, C., Daroowala, R., Goliath, R.T., Abrahams, F., Said-Hartley, Q., Allwood, B.W., Hsiao, N.Y., Wilkinson, K.A., et al. (2021a). Relationship of SARS-CoV-2-specific CD4 response to COVID-19 severity and impact of HIV-1 and tuberculosis coinfection. J Clin Invest 131. http://dx.doi.org/10.1172/JCI149125.

Riou, C., Keeton, R., Moyo-Gwete, T., Hermanus, T., Kgagudi, P., Baguma, R., Tegally, H., Doolabh, D., Iranzadeh, A., Tyers, L., et al. (2021b). Loss of recognition of SARS-CoV-2 B.1.351 variant spike epitopes but overall preservation of T cell immunity. medRxiv. http://dx.doi.org/10.1101/2021.06.03.21258307.

Robbiani, D.F., Gaebler, C., Muecksch, F., Lorenzi, J.C., Wang, Z., Cho, A., Agudelo, M., Barnes, C.O., Gazumyan, A., and Finkin, S. (2020). Convergent antibody responses to SARS-CoV-2 in convalescent individuals. Nature 584, 437–442. http://dx.doi.org/10.1038/s41586-020-2456-9.

Rydyznski Moderbacher, C., Ramirez, S.I., Dan, J.M., Grifoni, A., Hastie, K.M., Weiskopf, D., Belanger, S., Abbott, R.K., Kim, C., Choi, J., et al. (2020). Antigen-Specific Adaptive Immunity to SARS-CoV-2 in Acute COVID-19 and Associations with Age and Disease Severity. Cell 183, 996–1012 e1019. http://dx.doi.org/10.1016/j.cell.2020.09.038.

Saadat, S., Tehrani, Z.R., Logue, J., Newman, M., Frieman, M.B., Harris, A.D., and Sajadi, M.M. (2021). Binding and neutralization antibody titers after a single vaccine dose in health care workers previously infected with SARS-CoV-2. JAMA 325, 1467–1469. http://dx.doi.org/10.1001/jama.2021.3341.

Sadoff, J., Gray, G., Vandebosch, A., Cárdenas, V., Shukarev, G., Grinsztejn, B., Goepfert, P.A., Truyers, C., Fennema, H., and Spiessens, B. (2021). Safety and efficacy of single-dose Ad26. COV2. S vaccine against Covid-19. New England Journal of Medicine 384, 2187–2201. http://dx.doi.org/10.1056/NEJMoa2101544.

Sattler, A., Angermair, S., Stockmann, H., Heim, K.M., Khadzhynov, D., Treskatsch, S., Halleck, F., Kreis, M.E., and Kotsch, K. (2020). SARS-CoV-2-specific T cell responses and correlations with COVID-19 patient predisposition. J Clin Invest 130, 6477–6489. http://dx.doi.org/10.1172/JCI140965.

Schafer, A., Muecksch, F., Lorenzi, J.C.C., Leist, S.R., Cipolla, M., Bournazos, S., Schmidt, F., Maison, R.M., Gazumyan, A., Martinez, D.R., et al. (2021). Antibody potency, effector function, and combinations in protection and therapy for SARS-CoV-2 infection in vivo. J Exp Med 218. http://dx.doi.org/10.1084/jem.20201993.

Stamatatos, L., Czartoski, J., Wan, Y.H., Homad, L.J., Rubin, V., Glantz, H., Neradilek, M., Seydoux, E., Jennewein, M.F., MacCamy, A.J., et al. (2021). mRNA vaccination boosts cross-variant neutralizing antibodies elicited by SARS-CoV-2 infection. Science. http://dx.doi.org/10.1126/science.abg9175.

Stephenson, K.E., Le Gars, M., Sadoff, J., de Groot, A.M., Heerwegh, D., Truyers, C., Atyeo, C., Loos, C., Chandrashekar, A., and McMahan, K. (2021). Immunogenicity of the Ad26. COV2. S Vaccine for COVID-19. JAMA 325, 1535–1544. http://dx.doi.org/10.1001/jama.2021.3645.

Sykes, W., Mhlanga, L., Swanevelder, R., Glatt, T.N., Grebe, E., Coleman, C., Pieterson, N., Cable, R., Welte, A., and van den Berg, K. (2021). Prevalence of anti-SARS-CoV-2 antibodies among blood donors in Northern Cape, KwaZulu-Natal, Eastern Cape, and Free State provinces of South Africa in January 2021. Research Square. http://dx.doi.org/10.21203/rs.3.rs-233375/v1.

Takuva, S., Takalani, A., Garrett, N., Goga, A., Peter, J., Louw, V., Opie, J., Jacobson, B., Sanne, I., and Gail-Bekker, L. (2021). Thromboembolic Events in the South African Ad26. COV2. S Vaccine Study. New England Journal of Medicine 385, 570–571. http://dx.doi.org/10.1056/NEJMc2107920.

Tarke, A., Sidney, J., Methot, N., Yu, E.D., Zhang, Y., Dan, J.M., Goodwin, B., Rubiro, P., Sutherland, A., Wang, E., et al. (2021). Impact of SARS-CoV-2 variants on the total CD4(+) and CD8(+) T cell reactivity in infected or vaccinated individuals. Cell Rep Med, 100355. http://dx.doi.org/10.1016/j.xcrm.2021.100355.

Tauzin, A., Nayrac, M., Benlarbi, M., Gong, S.Y., Gasser, R., Beaudoin-Bussieres, G., Brassard, N., Laumaea, A., Vezina, D., Prevost, J., et al. (2021). A single dose of the SARS-CoV-2 vaccine BNT162b2 elicits Fc-mediated antibody effector functions and T cell responses. Cell Host & Microbe 29, 1137–1150 e1136. http://dx.doi.org/10.1016/j.chom.2021.06.001.

Tegally, H., Wilkinson, E., Giovanetti, M., Iranzadeh, A., Fonseca, V., Giandhari, J., Doolabh, D., Pillay, S., San, E.J., Msomi, N., et al. (2021). Detection of a SARS-CoV-2 variant of concern in South Africa. Nature 592, 438–443. http://dx.doi.org/10.1038/s41586-021-03402-9.

Vanshylla, K., Di Cristanziano, V., Kleipass, F., Dewald, F., Schommers, P., Gieselmann, L., Gruell, H., Schlotz, M., Ercanoglu, M.S., and Stumpf, R. (2021). Kinetics and correlates of the neutralizing antibody response to SARS-CoV-2 infection in humans. Cell Host & Microbe 29, 917-929.e914. http://dx.doi.org/10.1016/j.chom.2021.04.015.

Wang, P., Nair, M.S., Liu, L., Iketani, S., Luo, Y., Guo, Y., Wang, M., Yu, J., Zhang, B., Kwong, P.D., et al. (2021a). Antibody resistance of SARS-CoV-2 variants B.1.351 and B.1.1.7. Nature 593, 130–135. http://dx.doi.org/10.1038/s41586-021-03398-2.

Wang, Z., Muecksch, F., Schaefer-Babajew, D., Finkin, S., Viant, C., Gaebler, C., Hoffmann, H.-H., Barnes, C.O., Cipolla, M., and Ramos, V. (2021b). Naturally enhanced neutralizing breadth against SARS-CoV-2 one year after infection. Nature 595, 1–10. http://dx.doi.org/10.1038/s41586-021-03696-9.

Wibmer, C.K., Ayres, F., Hermanus, T., Madzivhandila, M., Kgagudi, P., Oosthuysen, B., Lambson, B.E., De Oliveira, T., Vermeulen, M., and Van der Berg, K. (2021). SARS-CoV-2 501Y. V2 escapes neutralization by South African COVID-19 donor plasma. Nature Medicine 27, 622–625. http://dx.doi.org/10.1038/s41591-021-01285-x.

Winkler, E.S., Gilchuk, P., Yu, J., Bailey, A.L., Chen, R.E., Chong, Z., Zost, S.J., Jang, H., Huang, Y., Allen, J.D., et al. (2021). Human neutralizing antibodies against SARS-CoV-2 require intact Fc effector functions for optimal therapeutic protection. Cell 184, 1804–1820 e1816. http://dx.doi.org/10.1016/j.cell.2021.02.026.

Zohar, T., Loos, C., Fischinger, S., Atyeo, C., Wang, C., Slein, M.D., Burke, J., Yu, J., Feldman, J., and Hauser, B.M. (2020). Compromised humoral functional evolution tracks with SARS-CoV-2 mortality. Cell 183, 1508-1519. e1512. http://dx.doi.org/10.1016/j.cell.2020.10.052.

