## Supplemental Data for "Prior infection with SARS-CoV-2 boosts and broadens Ad26.COV2.S immunogenicity in a variant dependent manner"

### Supplemental Table S1

**Supplemental Table 1: Clinical and demographic details of study participants relating to Figure 1**

| | No previous infection (n=20) | First wave infection (n=20) | Second wave infection (n=20) |
|---|---|---|---|
| **Demographic** | | | |
| Age (years)[b] | 48 [36-57] | 35 [30-38] | 36 [31-44] |
| Gender M:F (% Female) | 3:17 (85%) | 10:10 (50%) | 5:15 (75%) |
| Ethnicity | | | |
| Black | 1 (5%) | 5 (25%) | 7 (35%) |
| White | 6 (30%) | 9 (45%) | 5 (25%) |
| Mixed | 12 (60%) | 6 (30%) | 8 (40%) |
| Other | 1 (5%) | 0 (0%) | 0 (0%) |
| **Clinical** | | | |
| SARS-CoV-2 PCR positivity | 0% | 100% | 100% |
| Days after vaccination[b] | 30 [27-33] | 29 [28-33] | 28 [28-37] |
| Days from PCR+ test to vaccination[b] | N/A[c] | 232 [200-261] | 73 [54-82] |
| Disease severity | | | |
| WHO Scale 2 (mild)[d] | N/A | 20 (100%) | 20 (100%) |
| Comorbidities | | | |
| Asthma | 6 (30%) | 2 (10%) | 3 (15%) |
| Hypertension | 3 (20%) | 2 (10%) | 2 (10%) |
| Obesity | 2 (10%) | 2 (10%) | 1 (5%) |
| Diabetes mellitus | 2 (10%) | 2 (10%) | 1 (5%) |
| HIV | 0 (0%) | 0 (0%) | 0 (0%) |
| Other[e] | 1 (5%) | 1 (5%) | 2 (5%) |
| None | 8 (35%) | 13 (65%) | 12 (60%) |
| >1 comorbidity | 1 (5%) | 1 (5%) | 1 (5%) |

[a]Healthcare roles in the hospital included doctors (19), nurses (21), allied health professionals (11), administrative staff (5), cleaners (3), other (1); [b]median and interquartile range; [c]Not applicable; [d]World Health Organisation ordinal scale 2 (WHO Working Group on the Clinical Characterisation and Management of COVID-19 infection, 2020); [e]Other comorbidities not specified

### Supplemental Figure S1A

**A**

**Serological profiles - No prior infection**

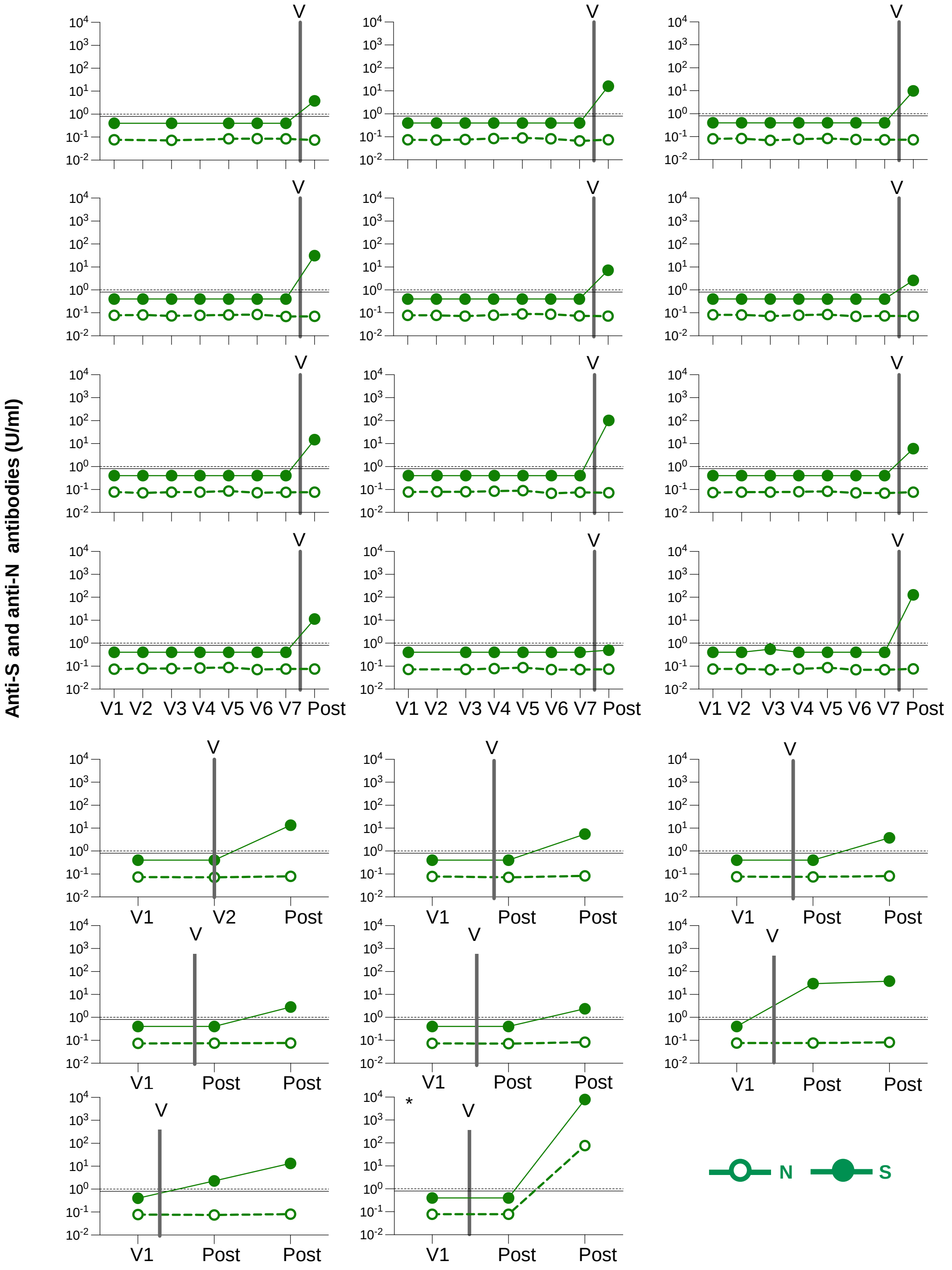

### Supplemental Figure S1B

**B**

**Serological profiles - First wave infection**

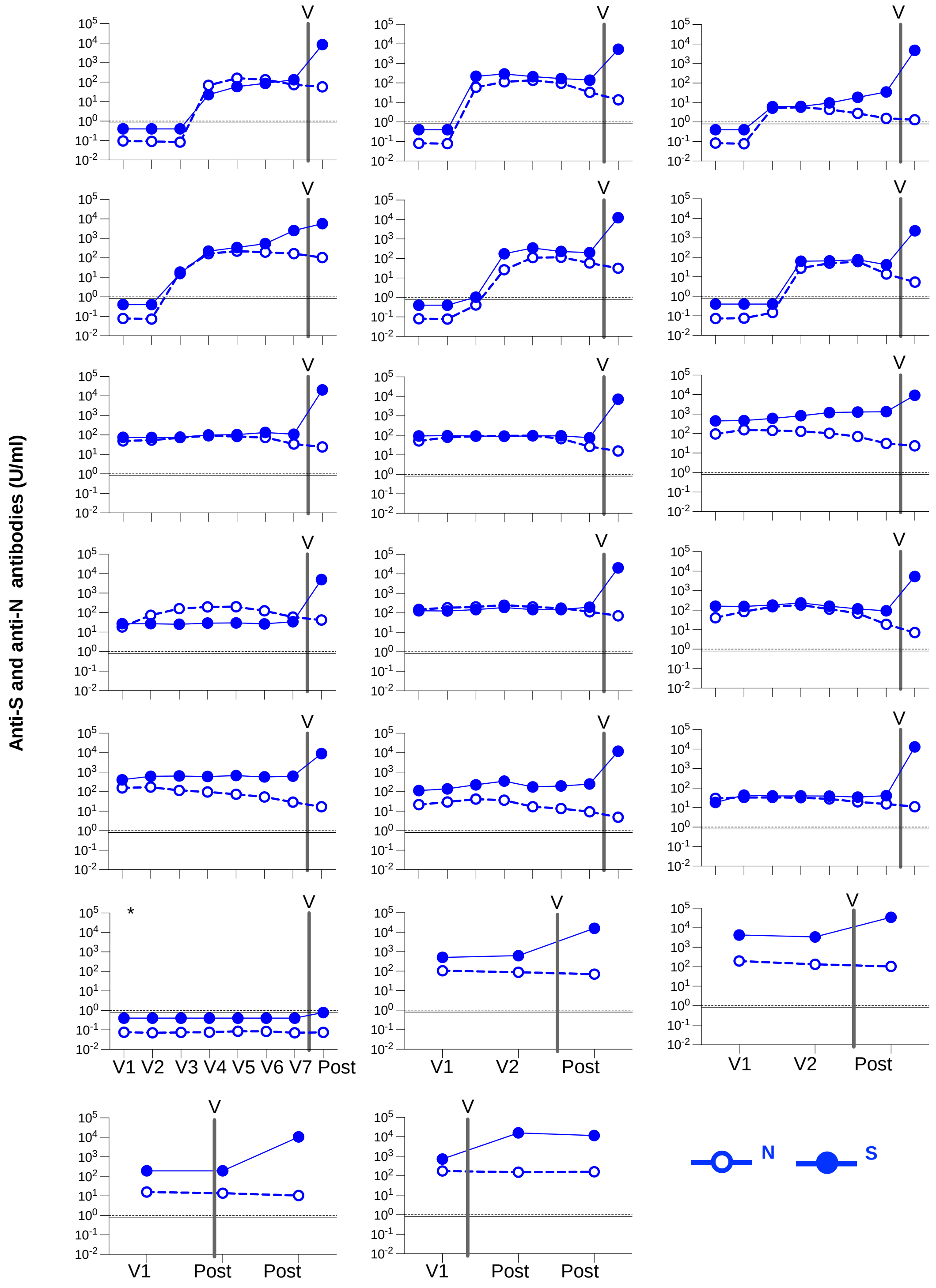

### Supplemental Figure S1C

C

**Serological profiles - Second wave infection**

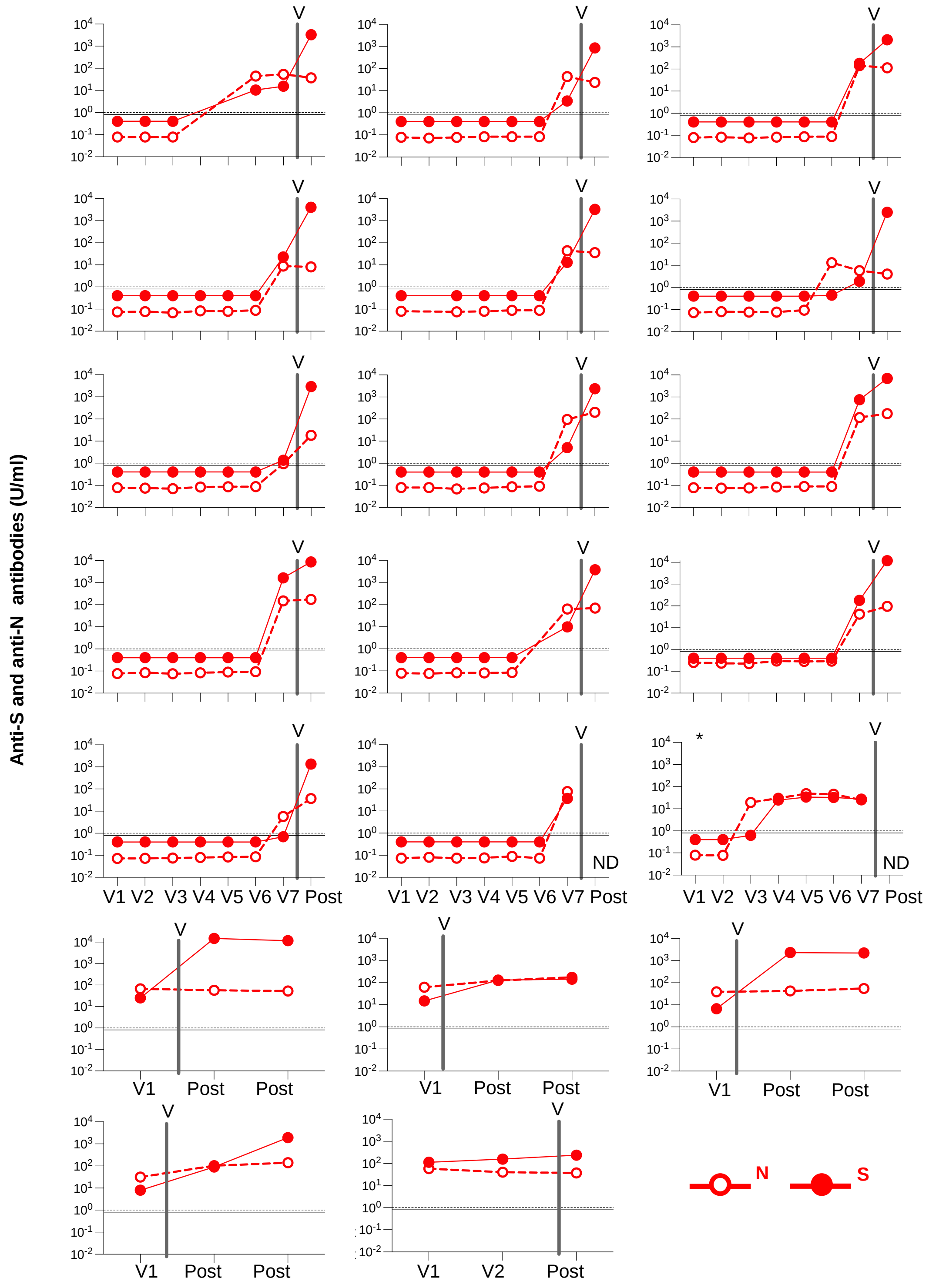


**Supplemental Figure 1: Serological profiles of study participants related to Figure 1**.
Spike and Nucleocapsid antibody profiles in A. No prior infection group; B. First wave infection; C. Second wave infection group. Serial serum samples were analysed from all available study visits prior to vaccination (3-8 samples per participant). Anti-spike (S; closed circles) and nucleocapsid (N; open circles) antibodies were measured by the Elecsys ECLIA system (Roche Diagnostics). The horizontal lines indicate the cut-off for a positive response (≥0.8 U/mL in the S assay, and ≥1.0 U/mL in the N assay). The vertical line with "v" indicates when vaccination took place. The asterisk indicates a potential breakthrough infection in A (both S and N antibodies increasing after vaccination); a serological non-responder despite a confirmed PCR test for SARS-CoV-2 in B; and a re-infection in C (a positive PCR in the second wave but serological evidence of infection in the first wave). The potential breakthrough and re-infection participants were excluded from further study. In B, 9 participants with the longer observation period were infected prior to the baseline sample (median 42 days, IQR 27-44) in July/August 2020.

### Supplementary Figure S2

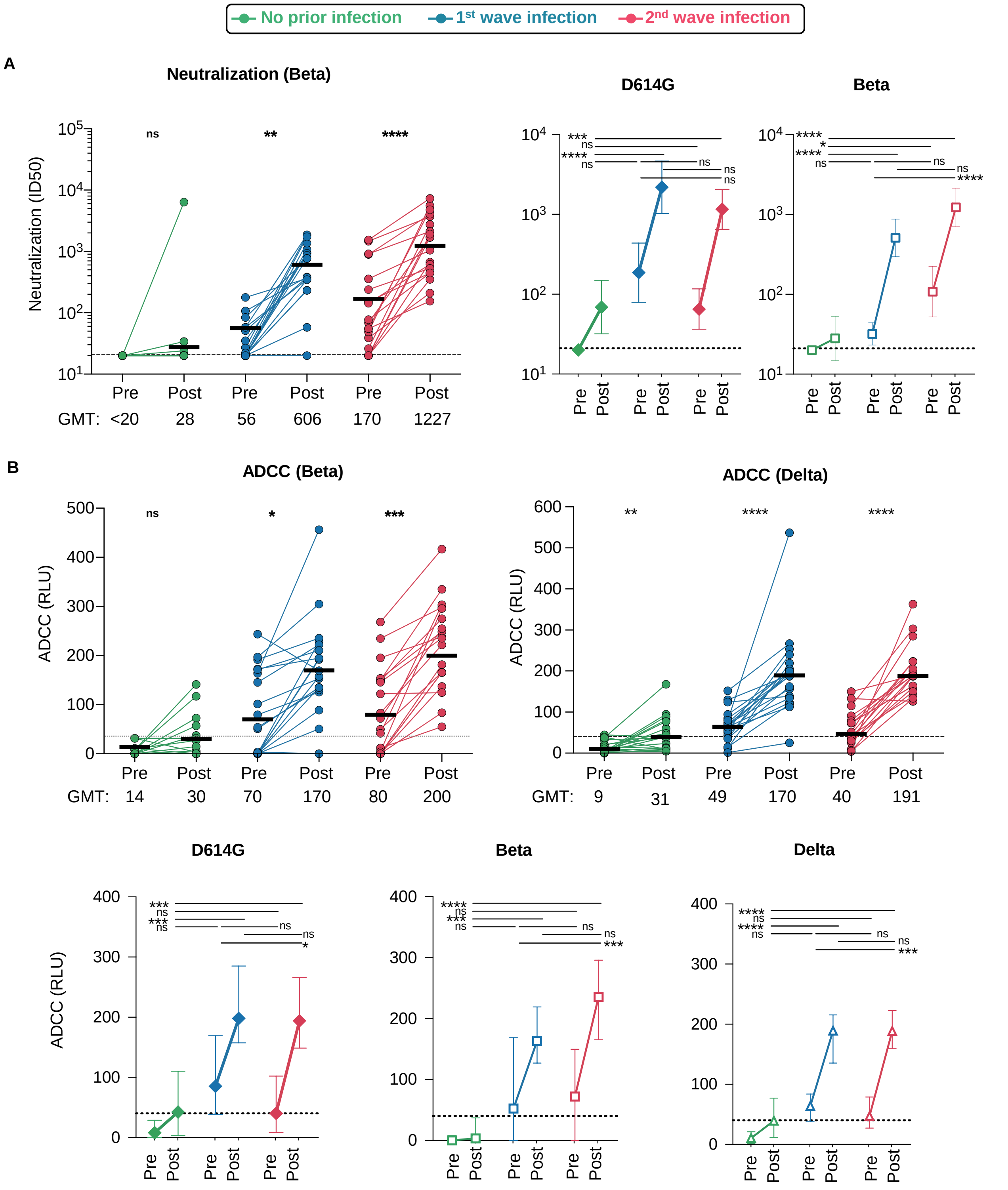


**Supplemental Figure 2: Neutralization and ADCC activity pre and post vaccination relating to Figure 2 and 3**
A. Neutralization of the SARS-CoV-2 Beta pseudovirus by plasma pre- and post-vaccination from participants with no prior infection (green, n=19) and those infected in the first (blue, n=20) and second waves (red, n=19). Neutralization is reflected as an ID50 titer. The threshold for positivity is indicated by a dotted line and GMT indicated below the graph and as bold black bars. Significance between pre and post vaccination was calculated by the Wilcoxon test. GMT pre and post vaccination are represented against D614G and Beta for each group, with significant represented by a Kruskal-Wallis test with Tukey correction. B. ADCC activity pre and post vaccination against Beta and Delta are shown as relative light units (RLU) with GMT represented below graphs and as before-after plots against D614G, Beta and Delta. * denotes p<0.05, ** p<0.01, *** p<0.001, ****<0.0001 ns, non significant. Experiments were performed in duplicate with the average value shown.

### Supplementary Figure S3

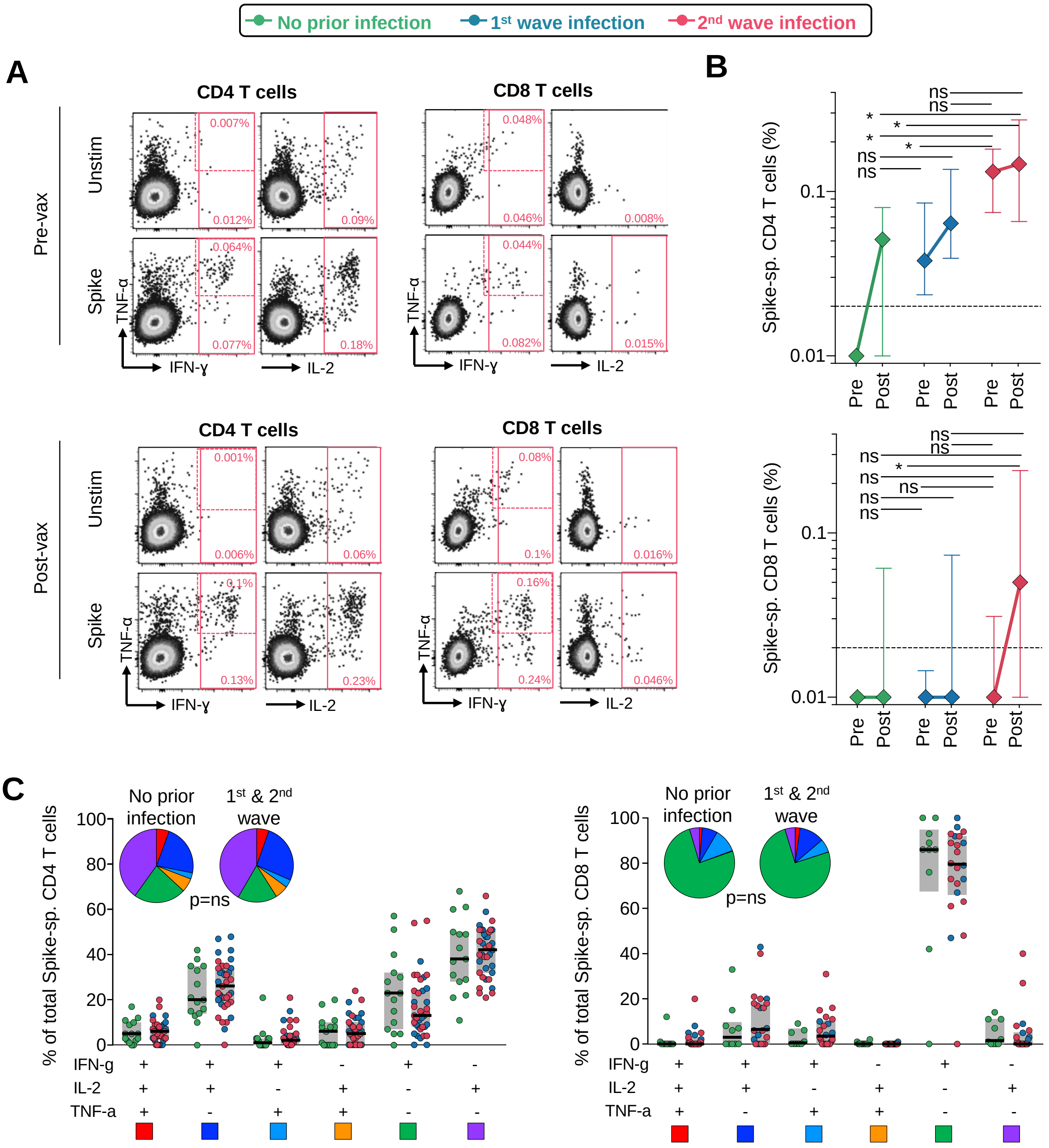


**Supplemental Figure 3**: **Analysis of T cell responses after Ad26.COV2.S vaccination relating to Figure 4.**
A. Representative flow cytometry plots of CD4 and CD8 T cell cytokine responses (IFN-g, TNF-a and IL-2) in response to a pool of spike peptides, with the unstimulated control shown. The pre- and post-vaccination plots are shown, from one second wave participant. T cell responses were calculated from boolean gates of all cytokines and the background (unstimulated sample) was subtracted. The single TNF-a-producing subset was excluded due to high background responses. B. Summary of median frequencies of cytokine-producing spike-specific CD4 and CD8 T cells, in those with no prior infection (green, n=19), infection in the first wave (blue, n=20), and infection in the second wave (red, n=19). Symbols represent medians and error bars IQR. Statistical comparisons between groups were performed with the Kruskal Wallis test with Dunn's multiple comparisons test. C. Polyfunctional analysis of CD4 and CD8 T cell responses post-vaccination. Comparison of the polyfunctional profile of spike-specific CD4 T cells (left panel) and CD8 T cells (right panel) in those without prior infection and those previously infected (first and second wave plotted together). Data are expressed as the proportion of each cytokine combination of the total response for each individual. The median and IQR are shown. Each response pattern is color-coded, and summarized in the pie charts. * denotes $p<0.05$, ns = non-significant.
